# Randomized controlled trials do not support efficacy of any of the tested doses of fluvoxamine in prevention of disease progression in adults with incipient non-severe COVID-19 disease: a case-study systematic review and meta-analysis

**DOI:** 10.64898/2026.04.01.26349972

**Authors:** Vladimir Trkulja

## Abstract

**Background:** Recent meta-analyses of randomized controlled trials (RCTs) claimed efficacy of higher-dose fluvoxamine (2 x 100 mg/day, as opposed to 2 x 50 mg/day) in prevention of disease deterioration in adults with mild - moderate COVID-19 disease.

**Objectives:** Investigate whether such claims are supported by the data.

**Methods:** Systematic review and meta-analysis of RCTs evaluating higher-dose fluvoxamine in this indication.

**Results:** Seven studies declared as RCTs were identified, one of which was severely biased (open-label, non-standardized and unreported standard of care as a control), and eventually ended as non-randomized (huge attrition). Composite endpoints of deterioration in the 6 included placebo-controlled trials contained elements susceptible to error and bias. Three trials were small (<100 patients/arm), three were larger (270 - 750 patients/arm). Deaths and need for mechanical ventilation were sporadic and observed in only one trial. Hospitalizations were also sporadic in 5/6 trials. Frequentist methods generally appropriate for random-effects analysis of low number of trials with rare outcomes (generalized linear mixed models, beta-binomial or binomial-normal) greatly underestimated heterogeneity, but still did not document benefits regarding the composite endpoints or hospitalizations. Bayesian hierarchical models revealed huge heterogeneity and indicated no benefit regarding: (i) composites of deterioration, large trials OR = 0.78 (95% CrI 0.55 - 1.21); multiplicity corrected OR = 0.87 (0.64 - 1.21); (ii) hospitalizations, small trials OR = 0.88 (0.45 - 1.72); large trials OR = 0.94 (0.52 - 1.75); all trials OR = 0.81 (0.47 - 1.43). Heterogeneity was unlikely due to clinical particulars (vaccination status, treatment duration, time horizon), and more likely due to unidentified bias.

**Conclusions:** RCTs do not support efficacy of higher-dose fluvoxamine in prevention of disease deterioration in adults with mild - moderate COVID-19 disease.

## Introduction

In April 2022, the United States Food and Drug Administration (FDA) declined a request for emergency use authorization of oral fluvoxamine for prevention of disease deterioration in adults with early mild-to-moderate COVID-19 disease (1). The decision was based on four randomized double-blind placebo-controlled trials (RCTs) conducted in unvaccinated adults: Stop COVID (2), Stop COVID 2 (3), Together (4), and Covid-OUT (5). Since then, additional RCTs have been published—most notably the Activ 6 trials (6,7)—which likewise failed to demonstrate its efficacy. Despite that, several relatively recent meta-analyses have suggested that higher-dose fluvoxamine—typically 2 × 100 mg/day—may be effective, in contrast to lower-dose protocols (i.e., 2 × 50 mg/day) (8,9). Given the current characteristics of SARS-CoV-2 and availability of effective vaccines and antiviral therapies, the clinical relevance of fluvoxamine in this indication has been diminished. Nevertheless, fluvoxamine remains easily accessible, and such claims may still encourage its inappropriate use. To clarify why these assertions are not substantiated by the available evidence—despite being based on meta-analyses of RCTs or studies labeled as such—the present work examines key aspects of the design and conduct of these trials that are essential for interpretation of their individual findings and for the validity of corresponding meta-analytic estimates.

## Methods

### Inclusion criteria and study identification

Eligible were published full-text reports (any language) of RCTs conducted in adults with confirmed early mild-to-moderate COVID-19 (Population) who were randomized to receive fluvoxamine at any dose or regimen exceeding 2 × 50 mg/day (claimed “not effective” (8,9)) (Intervention), with a control condition (Comparison) that permitted estimation of fluvoxamine’s effect, and which reported outcomes (Outcome) reflecting either (i) disease deterioration (e.g., hypoxemia/need for oxygen therapy, healthcare utilization such as hospitalization or urgent care visits, assisted ventilation, death, or any combination thereof) or (ii) symptom resolution (i.e., recovery).

To identify eligible primary studies, reference lists of the meta-analyses that motivated the present work (8,9) were searched, and electronic searches were conducted in PubMed/MEDLINE and the Cochrane Central Register of Controlled Trials using terms (SARS-CoV-2 OR COVID-19) AND fluvoxamine, with limits on publication date (January 1, 2020, to January 20, 2026) and study type (randomized controlled trial, clinical trial, systematic review, meta-analysis).

### Risk of bias

All studies included in the present assessment had been incorporated into at least one prior systematic review reporting efficacy of “higher-dose” fluvoxamine and had undergone formal risk-of-bias assessment (8,9). However, assessing risk of bias in RCTs is inherently complex and requires assessors to be specifically trained. In practice, such training is commonly not ensured, and assigned risk-of-bias ratings may vary substantially even among trained assessors (10,11). Post hoc appraisal of such judgments is limited by the fact that published papers rarely provide justification for the assigned ratings (e.g., 8,9). Consequently, it is often impossible to determine whether the original evaluations were appropriate and supported. Instead of repeating such assessments, the present work discusses key aspects of the design and conduct of each primary study, drawing on information available in the published reports and study protocols.

### Statistical considerations

Key methodological aspects typically scrutinized in pivotal trials of pharmacological therapeutics include: rationale for the sample size (expected baseline risk, anticipated effect size, and desired statistical power); details of the randomization process (e.g., concealment of the randomization list, allocation ratios, and stratified randomization procedures); prespecified analytical approaches for primary/secondary outcomes; and procedures for controlling the type 1 error rate (FWER). Each included trial was evaluated from this perspective.

Assigning a specific risk-of-bias level ideally requires judging the potential magnitude of bias that could have influenced the estimated treatment effect. In some cases, this may also require adjusting estimates to account for such biases (12). Accordingly, each study was examined with these considerations in mind: the main reported outcomes are summarized, and alternative effect estimates are presented when—given the limitations of the published reports—those alternatives appeared more likely to approximate the underlying treatment effect than the originally reported measures.

### Meta-analysis methods

Where appropriate, published and “alternative” (corrected for likely bias/chance) outcomes were meta-analyzed using Bayesian and frequentist methods appropriate for small numbers of heterogeneous trials with rare outcomes. All analyses were performed in R Language and Environment for Statistical Computing (R Core Team 2025) using packages *glmmTMB* (13), *metafor* (14) and *meta* (15) (frequentist beta-binomial, binomial-normal, and inverse-variance models, respectively), and *brms* (16) (Bayesian hierarchical models).

## Results

### Eligible studies

Of the 34 initial records, 23 were of inadequate design (Figure 1). Among the 11 full-text reports, three referred to lower fluvoxamine doses (2 × 50 mg/day)—Covid-OUT (5), Activ 6 with lower dose (7), and a smaller trial in Thailand (17). One additional study (18) compared fluvoxamine + inhaled budesonide to double placebo (Figure 1). Thus, seven studies declared as RCTs were included in the present review: Stop COVID (2), Stop COVID 2 (3), Together (4) [and study protocol (19)], a trial in Seoul, South Korea (20), Activ 6 (higher-dose cohort) (6), a single-center trial in Egypt (21), and a multicenter trial across several regions in Thailand (22) (Table 1).

**Table 1.**
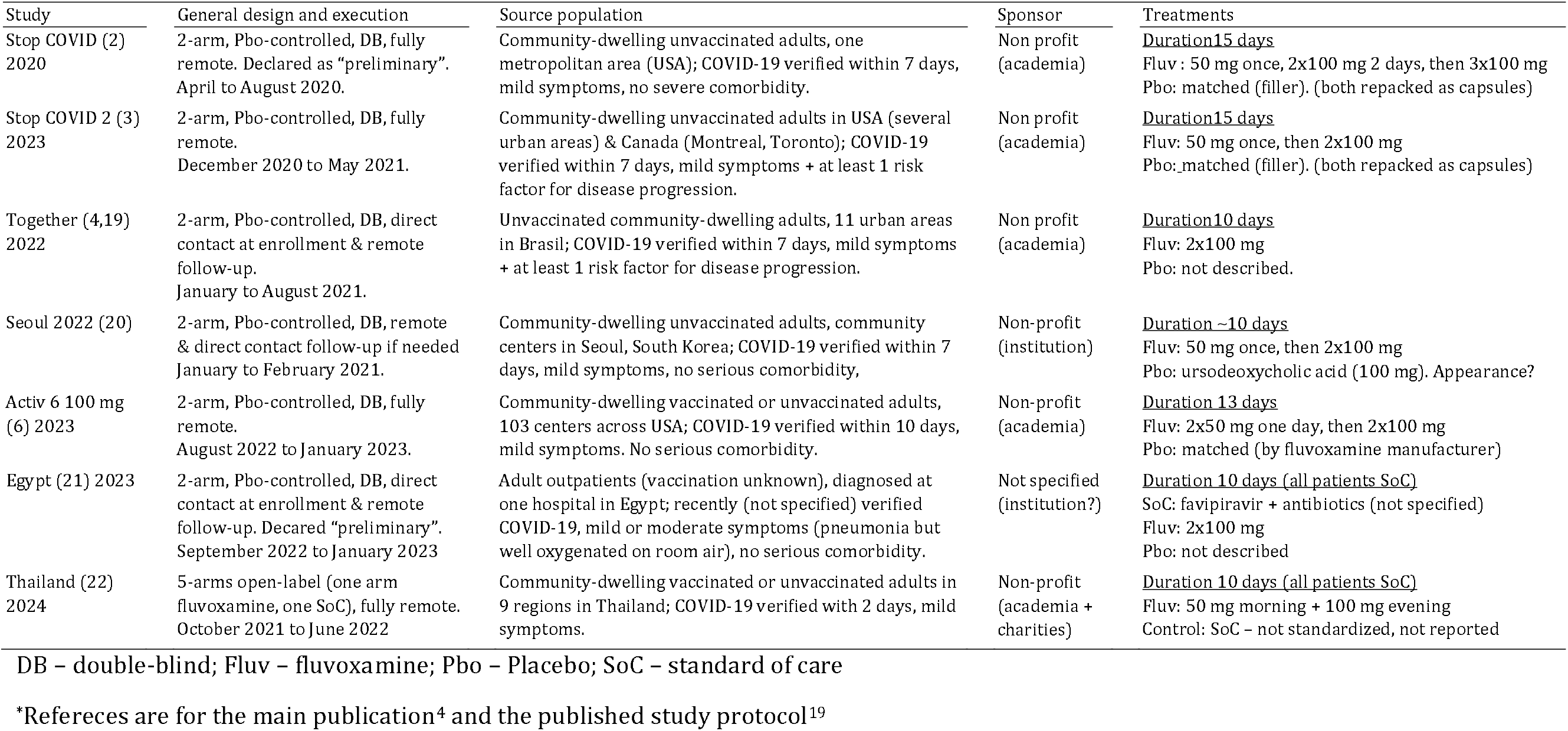
General characteristics of randomized trials of fluvoxamine for prevention of disease progression in adults with COVID-19 disease.

**Figure 1.**
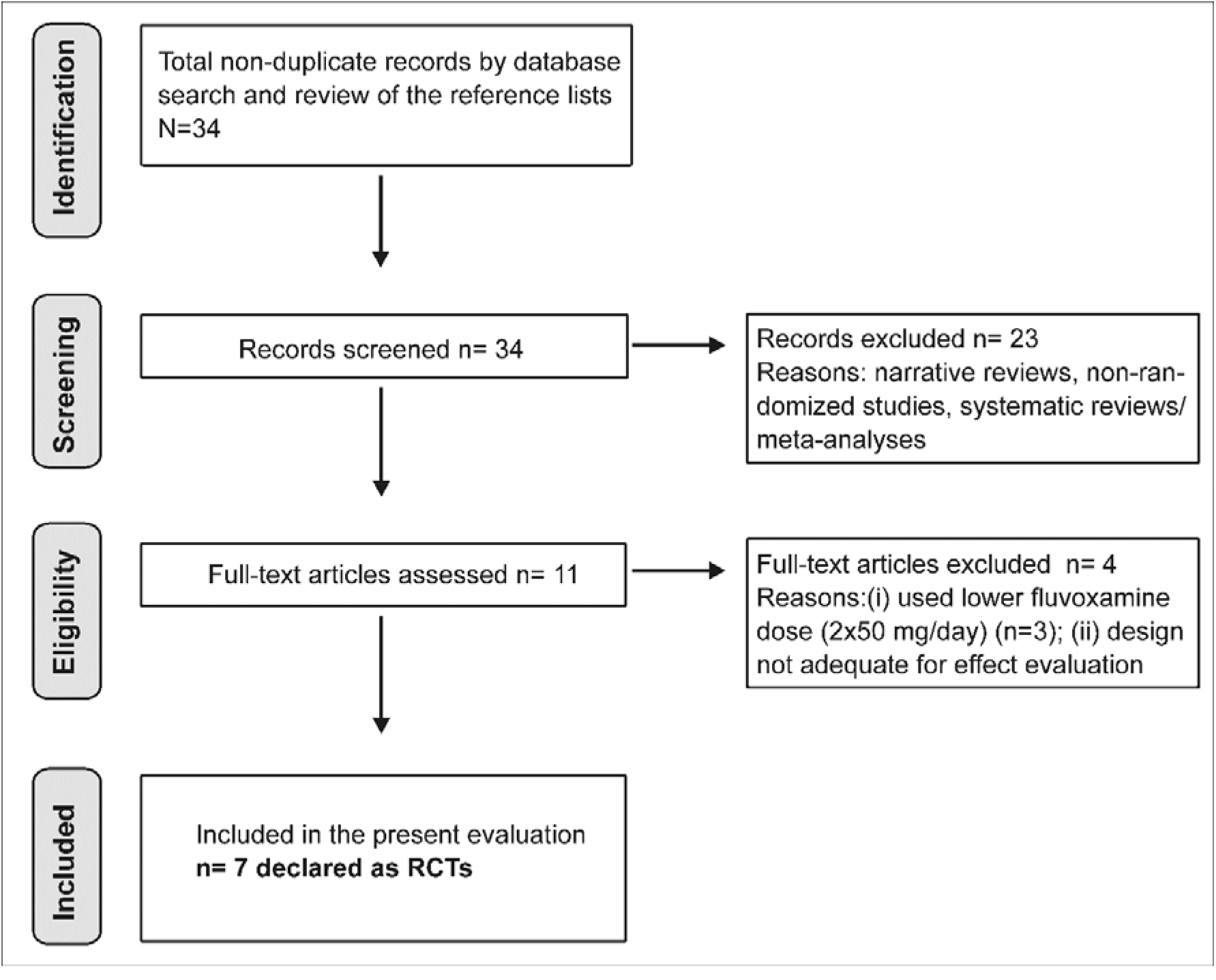
PRISMA flowchart. RCT – randomized controlled trial.

### General characteristics

The Thai trial (22) was open-label with standard of care (SoC) as the control, whereas other studies were declared as double-blind and placebo-controlled (Table 1). The trials were fully remote (2,3,6,22) or included direct contact at enrollment (4,21) or—if needed—at follow-up (20) (Table 1). Four trials included exclusively unvaccinated patients from large source populations in the USA or Canada (2,3), Brazil (4), or Seoul (20) (Table 1). Two trials recruited vaccinated and unvaccinated patients from large source populations in the USA (6) or Thailand (22), whereas one single-center study in Egypt (21) provided no information about vaccination status (Table 1). Patients had to be free of serious comorbidities, although in the Stop COVID 2 (3) and Together (4,19) trials, at least one established risk factor for disease progression was required (Table 1). Treatment was typically commenced within 7 days since diagnosis, with slight variation across trials (Table 1). Treatment duration was similar across trials (10–15 days) (Table 1).

Fluvoxamine doses—typically 2 × 100 mg/day (3,4,6,20,21), or 3 × 100 mg/day in one (2), and 50 mg (morning) + 100 mg (evening)/day in another (22) trial (Table 1)—may be considered a single treatment modality based on non-clinical pharmacological data (23,24). In the Stop COVID trials (2,3), fluvoxamine was repackaged into soft gelatin capsules matching placebo capsules, a modification unlikely to influence clinical outcomes (Table 1). In the Together trial (4,19), placebo was not described. In the Seoul trial (20), 100 mg tablets of ursodeoxycholic acid served as placebo (Table 1). It is uncertain how well it matched fluvoxamine tablets by appearance. Moreover, at least theoretically, ursodeoxycholic acid could affect COVID-19 outcomes through actions on gut microbiota and liver function (25) (Table 1). In the Egyptian trial (21), placebo was not described, leaving some uncertainty about its adequacy. Additionally, concomitant SoC treatments were specified only as favipiravir and antibiotics, without further detail (Table 1) (21). In the Thai trial (22), the control arm could vary from no antiviral treatment to several different treatments: it was not standardized and was not reported (Table 1).

### Outcome definitions, randomization, sample size and other statistical considerations

All trials planned multiple efficacy outcomes (Table 2). Composite measures of disease deterioration were included in all trials, but their definitions and time horizons differed (Table 2). When “oxygen desaturation” (via remote pulse oximetry) or remotely assessed dyspnea were components of these composites—as in the Stop COVID trials (2,3), the Seoul trial (20), and the Egyptian trial (21)—both measurement error and bias were possible due to the subjectivity of dyspnea ratings and the known limitations of pulse oximetry (26). In the Together trial (4), the validity of “≥6 hours of emergency room observation” as a proxy for hospitalization has been questioned (1), and this outcome is inherently vulnerable to subjective decision-making and organizational factors within the health-care system in which the trial was conducted. By contrast, the composite outcome in Activ 6 (6) consisted of unambiguous clinical events, and the severity scale used in the Thai trial (22) was likewise clearly defined (Table 2). Measures of disease resolution were planned as the primary outcome in Activ 6 (6) (28-day horizon) and as one of many secondary outcomes in Together (4) (14-day horizon) (Table 2).

**Table 2.** Details on outcome definitions, randomization, planned analyses and other statistical considerations.

| Study | Primary & other outcomes | Randomization | Statistical considerations | Planned analysis | Planned sample | Reported |
| --- | --- | --- | --- | --- | --- | --- |
| Stop COVID (2) 2020 | <u>Primary.</u> Deterioration day 15: (i) dyspnea or hospitalization + (ii) SatO<sub>2</sub> <92% or supplemental O<sub>2</sub>. Dyspnea remotely, SatO<sub>2</sub> pulse oximetry. <u>Secondary.</u> WHO scale day 15(remote). <u>Other.</u> Hospitalizations, deaths. | Centralized 1:1, stratified by age (4 levels) and sex. | <u>Sample size</u> (i) 20% base risk, 5% with treatment (15% ARR / 4-fold RRR); (ii) alpha 0.05; (iii) power 80%. <u>FWER control.</u> All outcomes apart the primary are “exploratory”. | <u>Primary.</u> product-limit proportions. Adjustments not mentioned. <u>Secondary.</u> scores <u>Other.</u> Simple counts. | Planned 180 1:1<br>Randomized: Fluv n=92<br>Pbo n=89 | FAS<br>Fluv n=80<br>Pbo n=72 |
| Stop COVID 2 (3) 2023 | As in Stop COVID. | Centralized 1:1, stratified age (2 levels), sex, site. | <u>Sample size</u> (i) 20% base risk, 13% with treatment (7% ARR / 35% RRR); (ii) alpha 0.05; (iii) power 80%. FWER control. Secondary & other outcomes no inference. | <u>Primary.</u> Δ product-limit proportions stratified by age, sex & site. <u>Secondary.</u> Δ scores <u>Other.</u> Simple counts. | Planned 880 1:1<br>Stopped - futility at interim with: Fluv n=334<br>Pbo n= 336 | mITT<br>Fluv n=272<br>Pbo n=275 |
| Together (4,19) 2022 | <u>Primary.</u> Hospitalization / emergency room (ER) visit >6 hours to day 28. <u>Secondary.</u> All cause death; Covid-related death; all-cause hospitalization to day 28, clinical resolution to day 14, and many others. | Centralized 1:1, stratified by age (2 levels) & site | <u>Sample size</u> (i) 15% base risk, 37.5% RRR with treatment; (ii) alpha 0.05; (iii) power 80%, Three interim & final analysis for the primary outcome <u>FWER control.</u> None – no account for multiple looks & outcomes. | <u>Primary.</u> Bayesian logistic regression. Prior unclear, adjustments not prespecified. <u>Secondary.</u> Frequentist adjusted methods, but adjustments not prespecified. | Planned around 750 per arm<br>Randomized: Fluv n=741<br>Pbo n=756 | ITT<br>Fluv n=741<br>Pbo n=756 |
| Seoul 2022 (20) | <u>Primary.</u> Deterioration day 30: (i) SatO<sub>2</sub> <94% or (ii) require O<sub>2</sub> or (iii) dyspnea; or (iv) WHO category ≥4. Dyspnea remotely, SatO<sub>2</sub> pulse oximetry. <u>Secondary.</u> Elements of the primary. | 1:1, stratified by age (4 levels) | Sample size (i) 10% base risk, 4-fold RRR with treatment; (ii) alpha 0.05; (iii) power 80%. Stopped early after only 52 patients had been enrolled. | Unadjusted differences in proportions. | Planned 400 1:1<br>Stopped due to lack of patients at Fluv n=26<br>Pbo n=26 | ITT<br>Fluv n=26<br>Pbo n=26 |
| Activ 6 100 mg (6) 2023 | <u>Primary.</u> Time to recovery to day 28. <u>Secondary.</u> (i) Time to health care utilization to day 28 (hospitalization, urgent care, ER visit or death); (ii) hospitalization or death to day 28; (iii) deaths; (iv) cumulative hospitalizations. | Centralized 1:1 | <u>Sample size</u> (i) 20% increase in risk of recovery; (ii) alpha 0.05; (iii) power 80%. Three interim & final analysis. <u>FWER control.</u> Skeptical prior (N(0.0.1)) at each for the primary. Other outcomes: no inference. | <u>Primary & secondary</u> - adjusted for age, sex, time-to-treatment, calendar time, vaccination, region, center, and baseline symptom severity. | Planned 1200 1:1<br>Randomized: Fluv n=601<br>Pbo n=607 | mITT<br>Fluv n=589<br>Pbo n=586 |
| Egypt (21) 2023 | <u>Primary.</u> Deterioration to day 10: SatO<sub>2</sub> <93% (method?)/Covid hospitalization. <u>Secondary.</u> Hospitalization, death. | Not described | No information on any statistical aspect (e.g., sample size, power, FWER) | No information on any data-analysis aspect. | No information. Mild+moderate enrolled. | Completers<br>Fluv n=37 + 51<br>Pbo n=35 + 39 |
| Thailand (22) 2024 | “Shifts” on a 6-point scale (not hospitalized, to hospitalized with or without O<sub>2</sub> supplementation and death) at days 9, 14, and 28 -assessed remotely. | Centralized 1:1<br>Stratified by age (4 levels) and sex. | <u>Sample size</u> (i) base risk of deterioration 3.2%, 0 with fluvoxamine; (ii) alpha 0.05; (iii) power 80%. <u>FWER control</u> - none. | Regression modelling of repeated ordinal data. Unclear planned adjustments. | Randomized<br>Fluv=374<br>SoC= 390 | Treated<br>Fluv n=162<br>SoC n=336<br>Not all times. |

ER – emergency room; FAS – full analysis set (received at least one dose of the assigned treatment); Fluv – fluvoxamine; FWER – familywise type 1 error rate; (m)ITT – (modified)
intent-to-treat set (mITT - received at least one dose of the assigned treatment); Pbo – placebo; SatO<sub>2</sub> – oxygen saturation; SoC – standard of care; WHO - World Health Organization

Randomization procedures were not described in the Egyptian trial (21), whereas other trials reported procedures that appeared adequate to prevent selection bias, typically involving stratification on one or two prognostic factors (Table 2).

In Stop COVID (2), sample size assumptions were based on an expected 20% baseline risk of deterioration and an implausibly large fluvoxamine effect (15% absolute risk reduction, ARR, corresponding to a fourfold relative risk reduction, RRR), using a product-limit estimator without adjustment for stratified randomization (Table 2). The analyzed full analysis set (FAS; individuals receiving at least one dose) was 15–20% smaller than the randomized sample (Table 2). Only the primary endpoint was designated for formal inference; no control of the family-wise error rate (FWER) was planned (2).

Stop COVID 2 (3) followed the same logic in all aspects except that the expected effect size was less optimistic (7% ARR) (Table 2), and stratification-variable adjustment was planned for the primary endpoint (Table 2). The trial was stopped for futility at interim analysis. The modified intent-to-treat set (mITT, defined as the FAS in Stop COVID) was around 20% smaller than the randomized sample (Table 2).

In the Together trial (4,19), sample size was planned on the expected baseline risk of 15% for deterioration, with a marked expected fluvoxamine effect of 37.5% RRR. The planned sample was randomized and analyzed (intent-to-treat, ITT) (Table 2). Three interim analyses were planned (19). The investigators argued that within a Bayesian framework, no adjustment akin to frequentist FWER control was necessary (19); however, empirical evidence shows that repeated interim looks increase the risk of false-positive findings, and regulatory standards require Bayesian approaches to incorporate safeguards equivalent to FWER control, such as skeptical priors (27). The Together trial protocol remained vague about the priors used for the primary endpoint (19), nor did it discuss FWER control for numerous secondary outcomes analyzed using frequentist methods. Covariate adjustment was planned for primary and secondary endpoints, but instead of pre-specifying covariates, the protocol stated that variables showing baseline imbalance would be included (19). Such post hoc covariate selection is methodologically unacceptable in a regulatory context, as it constitutes a form of “fishing.” The rationale for selecting “imbalanced” covariates misunderstands the purpose of randomization, which ensures exchangeability rather than perfect balance (28). Stratification creates correlation between treatment assignment and covariates; therefore, failing to adjust for stratification factors can bias estimates or inflate type I or type II error rates (29). Regulatory guidance requires adjusting for stratification factors and important prognostic covariates (30). The trial in Seoul (20) was stopped due to lack of patients when only 10% of the planned sample (26 per arm, ITT) were enrolled (Table 2). Given this circumstance, other considerations of its statistical properties are irrelevant.

In Activ 6 (6), the sample size was based on detecting a 20% improvement in time to recovery (primary endpoint). The mITT sample closely matched the randomized population (<5% discrepancy) (Table 2). Three interim and a final analysis were planned, each using a skeptical prior for ln(hazard ratio) (N(0, 0.1)), with adjustment for several prespecified covariates (Table 2). Covariate adjustment was also planned for the composite deterioration endpoint; however, a flat prior was used and no inferential interpretation was intended for secondary outcomes (Table 2).

In the Egyptian trial (21), no statistical rationale, planned analyses, or randomization details were reported. Only results from a small group of “completers” were provided, with no information about the originally randomized population, attrition, or reasons for dropout (Table 2).

In the Thai trial (22), sample size calculations assumed a 3.2% hospitalization risk in the control arm and an unrealistically optimistic 0% risk in the fluvoxamine arm (Table 2). Ultimately, only 162/374 patients randomized to fluvoxamine and 336/390 randomized to SoC received treatment, and even fewer had outcomes recorded at scheduled time points through day 28 (22).

### Reported study results and important considerations

In Stop COVID (2) (Table 3), bias and error were likely due to the elements of the primary endpoint and disregarded stratification. For hospitalization alone, the difference was smaller but still favored fluvoxamine: 0/80 vs. 4/72 (RD = –5.5%, 95% CI –13.5 to –0.8) (Table 3). However, with only 56% power to detect an effect of this magnitude, the probability that such a finding resulted from sampling variability was similar to the probability of observing a true effect. If the true rate with fluvoxamine were indeed zero, fluvoxamine would appear more effective than nirmatrelvir/ritonavir, a recommended therapy for this indication, which achieved a 1% event rate (hospitalization or all-cause death) in its pivotal trial (31). Assuming the true fluvoxamine rate was also 1%, a sample of 80 treated participants would yield a 38.1% probability of observing zero events and a 37.0% probability of observing exactly one event (Table 3). Thus, the apparent benefit—particularly the “zero” rate—was fragile and plausibly due to chance. Supporting this interpretation, the corresponding composite event rate in Stop COVID 2 was 4.8%, and hospitalizations occurred in 3.3% of participants (Table 3), despite both trials enrolling unvaccinated participants from the same source population (Table 1).

**Table 3.**
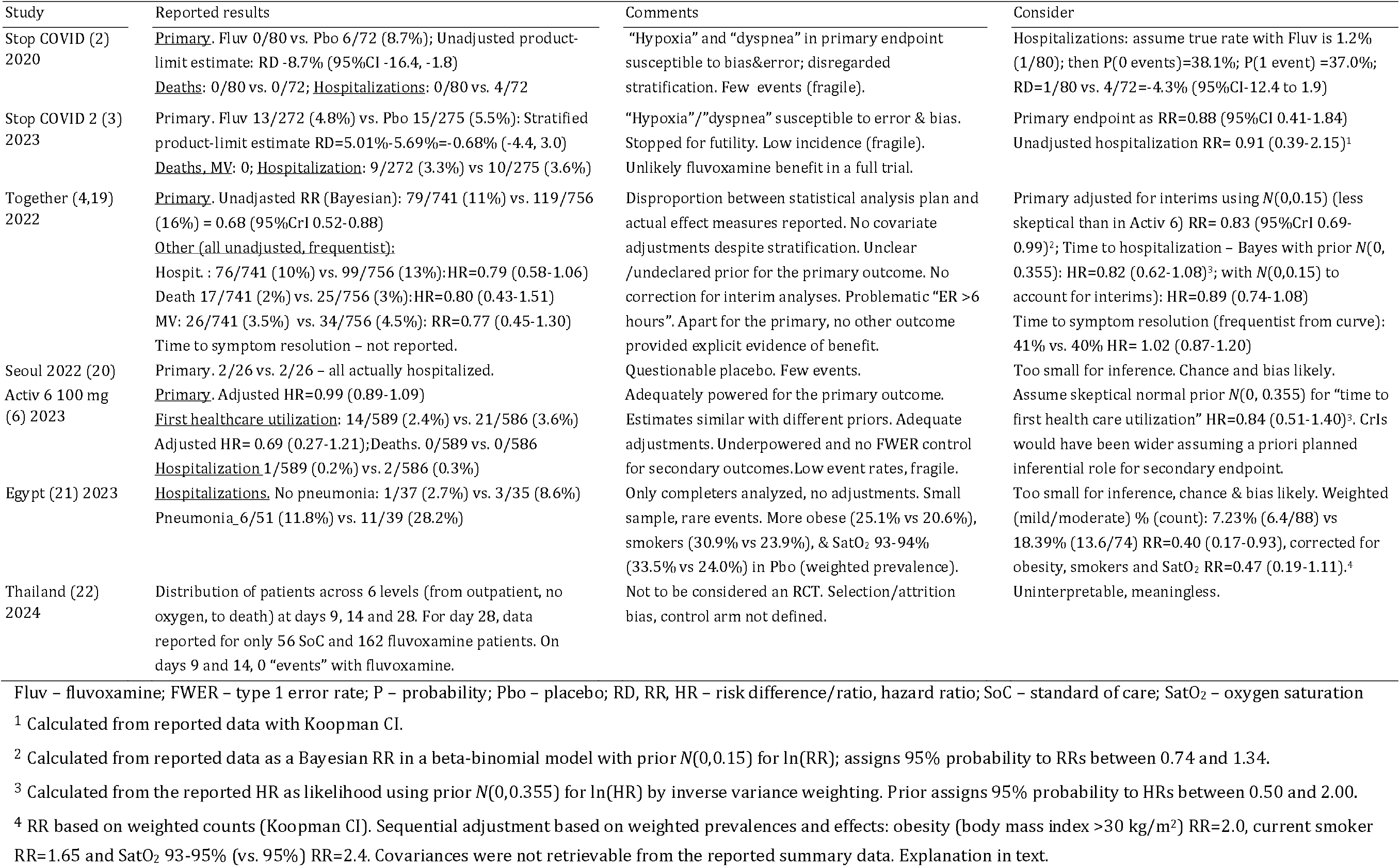
Reported trial results and related comments/considerations.

In Stop COVID 2 (3), the adjusted estimate for deterioration at the interim analysis showed no benefit of fluvoxamine (Table 3). Hospitalization rates were likewise low and similar between groups (Table 3). Given the low incidence and lack of signal, it is highly unlikely that continuation to full enrollment would have altered the conclusion.

The Together trial (4) suggested a marked effect of fluvoxamine regarding deterioration in unvaccinated patients: RR = 0.68 (95% CrI 0.52–0.88) (with considerably higher event rates for placebo (11%) and fluvoxamine (16%) than in the Stop COVID 2 trial) (Table 3). Several issues are noteworthy: (i) all estimates were unadjusted despite stratification; (ii) the primary analysis used a Bayesian beta-binomial model, but the prior was not specified and interim analyses were not incorporated. When applying a prior less skeptical than that used in Activ-6 (Table 2)—*N*(0, 0.15), which assigns 95% probability to RRs 0.74–1.35—the estimate becomes RR = 0.83 (95% CrI 0.69–0.99); (iii) the reported unadjusted estimate (frequentist) for hospitalization was HR = 0.79 (95% CI 0.58–1.06) (Table 3). Treating this as an unbiased estimate of deterioration risk and applying a moderately skeptical prior consistent with a priori equipoise (*N*(0, 0.355), assigning 95% probability to HRs 0.5–2.0) yields HR = 0.82 (95% CrI 0.62–1.08) (Table 3). Accounting for the multiple interim looks, a more skeptical prior (N(0, 0.15)) gives HR = 0.89 (95% CrI 0.74–1.08) (Table 3); (iv) time-to-resolution results—planned in the protocol (Table 2)—were not reported (4), but can be extracted from the published Kaplan–Meier curve (32), yielding HR = 1.02 (95% CI 0.87–1.20) (Table 3). With a control event rate of 40%, the study had approximately 80% post hoc power to detect HR = 1.20. Even allowing for an estimated 25% bias (in either direction), the corrected HR would still fall within approximately 0.95 to 1.05. The trial in Seoul (20) was only a “fragment” of a trial, too limited (and possibly biased) for inference. All four patients with the primary event were actually hospitalized (Table 3).

In Activ 6 (6), outcomes were analyzed as pre-specified (Table 2). No benefit was observed for time to recovery (Table 3). For the composite deterioration endpoint—analyzed with a flat prior due to its predefined non-inferential role—the estimate was HR = 0.69 (95% CrI 0.27–1.21) (Table 3). The number of events was low (2.4% vs. 3.6%), making the estimate fragile. If one were to impose inferential meaning using a moderately skeptical prior (*N*(0, 0.355)), the resulting estimate would be HR = 0.84 (95% CrI 0.51–1.40).

The Egyptian trial (21) reported only “completer” data, without describing randomization, blinding, or placebo adequacy (Table 2). Substantial baseline imbalances were present, and event numbers were very small (Table 3), making bias and chance highly likely explanations for the reported findings. Weighted proportions indicated apparent benefit, and an external adjustment for imbalances (33) in obesity, smoking, and baseline oxygen saturation (93–94% vs. ≥95%) could be applied using published risk ratios: approximately 2.0 for obesity (combined categories), 1.65 for current smoking (34), and 2.4 for prehospital oxygen saturation of 92–94% (35). The resulting adjusted estimate (RR = 0.47, 95% CI 0.19–1.11) (Table 3) remains plausibly biased and cannot support a conclusion of benefit.

The Thai trial (22) suffered from extreme discrepancies between randomized and treated participants (Table 2), as well as substantial additional attrition for outcome assessments (Table 3). Given these departures from protocol, combined with its open-label design and highly heterogeneous, unreported control treatments (Table 2), the study cannot be regarded as a valid randomized trial. Accordingly, its unadjusted results are uninterpretable.

### “Higher dose” fluvoxamine RCTs – what and how to meta-analyze, if anything?

A systematic review should filter the best available evidence to support an accurate estimate of a treatment effect. Because one can seldom be certain about biases in a primary study, different levels of certainty must be reflected in the assigned risk-of-bias ratings (36). Trials judged at high risk across standard domains typically report exaggerated effects compared with low-risk trials (37). Sensitivity analyses—such as excluding high-risk studies—may help identify the true treatment effect (38). Concordance between the “main” analyses (all studies) and “sensitivity” analyses (excluding high-risk studies) is often interpreted as supporting robustness (38).

However, when a study is explicitly biased, its contribution to accuracy is questionable: (i) such trials primarily estimate the bias itself, and one cannot know what the effect would have been without it; and (ii) effect estimates from low-risk studies are not adequate benchmarks— estimates from heterogeneous studies/populations pertain to different true effects. Thus, it is reasonable to disregard studies with explicit and severe bias rather than integrate them in pursuit of an unbiased pooled estimate. The Thai trial (22) meets this criterion and is therefore not considered further.

#### Acceleration of disease resolution

Both Activ-6 (6) and the Together trial (4) produced estimates closely around unity (Table 3). No meta-analysis is needed—homogeneous evidence of no clinically relevant fluvoxamine effect in unvaccinated and/or vaccinated patients is obvious.

#### Mortality and need for mechanical ventilation

Except in the Together trial (4), no deaths or mechanical ventilation events occurred (Table 3); hence a meta-analysis is meaningless. In Together, uncertainty was large due to sparse events: mortality 2% vs. 3% (95% CIs from 57% lower to 51% higher risk), and mechanical ventilation 3.5% vs. 4.5% (95% CIs from 55% lower to 30% higher risk) (Table 3). Hence, no reasonable evidence of benefit was observed.

#### Hospitalization and composite endpoints illustrating deterioration

Meta-analysis of clinically heterogeneous RCTs requires random-effects models and estimation of between-study variance (τ^2^). With few studies, τ^2^ estimates are often severely biased, and no estimator is ideal. This affects study weights, pooled effects, and prediction intervals (39), especially in meta-analyses of binary outcomes with low incidence or zero events (40). Multiple models have been proposed for such situations (41), but classical methods perform poorly—typically excluding zero-event studies or applying continuity corrections, both of which introduce bias (41). In the frequentist framework, generalized linear mixed models, beta-binomial models (42), and binomial-normal models (43) are preferred. Yet when heterogeneity is substantial and the number of trials small, frequentist methods often fail, typically estimating τ^2^ as exactly zero and producing falsely narrow confidence intervals (41). Bayesian hierarchical models handle sparse data more appropriately by directly modeling τ^2^ and propagating its uncertainty (41). In this context, the prior on τ^2^ is critical; weakly informative priors help constrain implausible heterogeneity (44). A further problem arises from pooling data across small and larger studies: (i) studies not sufficiently powered for a particular effect size may be considered “small” (45); (ii) small studies are more likely methodologically flawed and, when published, commonly report inflated effects (“small-study effect bias”) (45,46); (iii) with only a few studies, methods to approximate such bias are underpowered and not useful (45); (iv) small studies are more variable than larger ones. When combined, small trials distort τ^2^ estimation, giving them inappropriately high weight and diminishing the influence of larger trials (47). Thus, the assumption that “all available evidence” should be pooled to approach the truth is questionable, especially with infrequent binary outcomes (46). It is more prudent to analyze small and large trials separately.

Regarding “hospitalizations,” Figure 2 illustrates that—even assuming no bias—the three small trials—Stop COVID (2), Seoul (20) and the Egyptian (21) trial—produce statistical noise rather than meaningful information. A Bayesian model applied to the raw data demonstrates notable heterogeneity and wide prediction intervals, with the pooled estimate (OR = 0.88, 95% CrI 0.45– 1.72) spanning from markedly lower to markedly higher odds of hospitalization (Figure 2A). Frequentist models fail to meaningfully estimate τ^2^ (mean 0, CI up to 3.57), yielding unstable pooled estimates (Figure 2A). Using data “corrected” for random chance in Stop COVID (one event instead of none with fluvoxamine) and baseline imbalances in the Egyptian trial leads to the same conclusion: substantial heterogeneity and wide uncertainty under Bayesian models, and unestimable τ^2^ with very wide frequentist CIs (Figure 2B).

**Figure 2.**
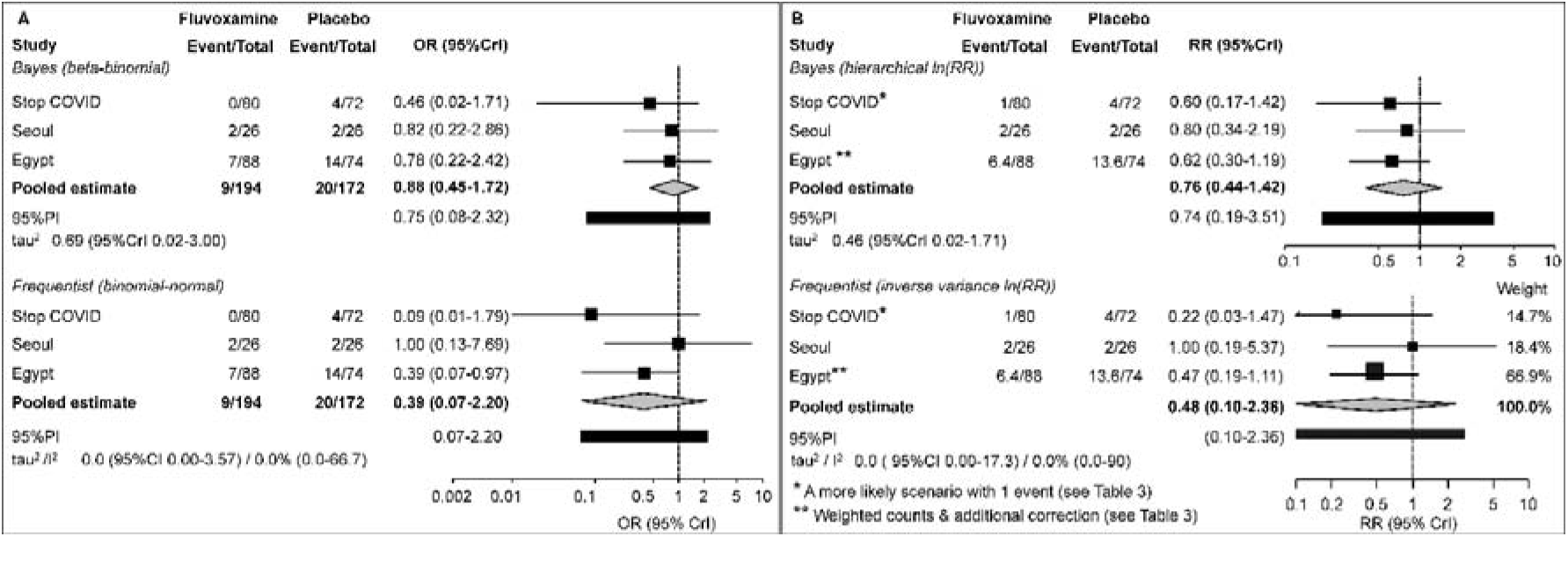
Random-effects meta-analysis of incidence of hospitalization in three small trials: Stop COVID (2), Seoul (20) and Egyptian (21) trial. The latter two trials only reported hospitalizations, whereas in Stop COVID, hospitalizations are devoid of error/bias of the other elements of the composite endpoint (Table 3). Meta-analyses were performed on reported data (A), and on “corrected” data (B) – with one event in the fluvoxamine arm in Stop COVID as a more realistic option than the reported zero events, and using weighted counts and correction for confounder imbalances in the Egyptian trial (Table 3). Bayesian and frequentist methods were employed. **A.** Data as reported. The Bayesian beta-binomial model used moderately informed skeptical prior for the treatment effect (*N*(0, 0.355) - fixed at 0 for ln(OR) with SD 0.355, assigns 95% probability to ORs 0.5-2.0, and weakly informative prior for τ^2^ (Cauchy (0, 0.5)). Frequentist beta-binomial model did not converge, hence binomial-normal conditional model with approximate likelihood and t-distribution was fitted. Bayesian study-level odds ratios are shrinkage estimates; frequentist odds ratios are model-derived estimates. **B**. Corrected data. Logarithms of precalculated relative risks were modelled. Bayesian model used the same priors as for the reported data. Study-level relative risks are shrinkage estimates. Frequentist model was a generic inverse variance model with restricted maximum likelihood estimator with Hartung-Knapp-Siddik-Jonkman and ad-hoc variance correction.

Considering the deterioration endpoints in the three larger trials, it appears more appropriate to use reported effect measures than raw counts per treatment arm (48): (i) in Stop COVID 2, stratified risk difference convertible to RR (Table 3); (ii) in Together, unadjusted RR from a Bayesian beta-binomial model with an apparently flat prior (Table 3); (iii) in Activ 6, adjusted hazard ratio with a flat prior; because events were rare (Table 3), it corresponds to RR (49). As discussed earlier, the Together estimate should be corrected for interim looks (to control false-positive risk), and the Activ 6 estimate should similarly be corrected because the statistical framework was explicitly non-inferential (Table 2, Table 3). Figure 3A shows meta-analysis of reported estimates, and Figure 3B shows meta-analysis of corrected estimates. For both types of data, Bayesian models indicate substantial heterogeneity with wide prediction intervals (Figure 3A,B). The estimate based on reported effects (RR = 0.78, 95% CrI 0.55–1.21) indicates from 45% lower to 21% higher risk of the event with fluvoxamine (Figure 3A), whereas the corrected estimate (RR = 0.87, 95% CrI 0.64–1.21) indicates from 36% lower to 21% higher risk (Figure 3B). Frequentist models again struggle to estimate τ^2^ (estimated as zero), producing unstable pooled RRs (0.70 and 0.83) with wide uncertainty (Figure 3A,B). Heterogeneity—rather than power or sample size—is the dominant problem. In this respect, it should be noted: (i) there were 1602 treated and 1617 control subjects (total N = 3219) in the three trials, with 155 events in the placebo arms (incidence 0.096); (ii) if one assumes three trials, each with a total of 1000 patients (3000 in total), and inconsistency index (I^2^) of 90% (corresponding to its upper 95% CI limit in Figure 3A) for the frequentist estimate OR = 0.70 (Figure 3A), there is still 83% probability to “place” the 95% CIs around the estimate to below 1.0. Obviously, heterogeneity and inconsistency were remarkable, and study characteristics (unvaccinated or mixed populations, timing and duration of treatment) (Table 1) do not suggest possible reasons. This estimate (Figure 3A) was markedly dominated by the Together trial. This suggests that idiosyncratic features of that trial—pertaining to its population, operational conduct, or outcome ascertainment—may explain its divergence. In particular, the primary composite included ≥6 hours of emergency-room observation, a subjective element potentially susceptible to differential misclassification and ascertainment bias, especially if the assigned treatment could be “unblinded.” The problem of “unblinding” based on recognized biological effects is known for antidepressants in the treatment of depression, where it may affect patient responses (by affecting expectations, i.e., placebo or nocebo effects) (50), but it might also affect decisions of the investigators, particularly those overtly enthusiastic about the treatment. Figure 4 further dissects hospitalization and non-hospitalization components. Stop COVID 2 and Activ 6 show balanced distributions of these components between arms, whereas Together shows an anomalously low proportion of non-hospitalization events in the fluvoxamine group (3/79) but high in the placebo group (20/119). Bayesian and frequentist models again cannot establish a benefit of fluvoxamine, either regarding hospitalizations (Figure 4A) or non-hospitalization events (Figure 4B).

**Figure 3.**
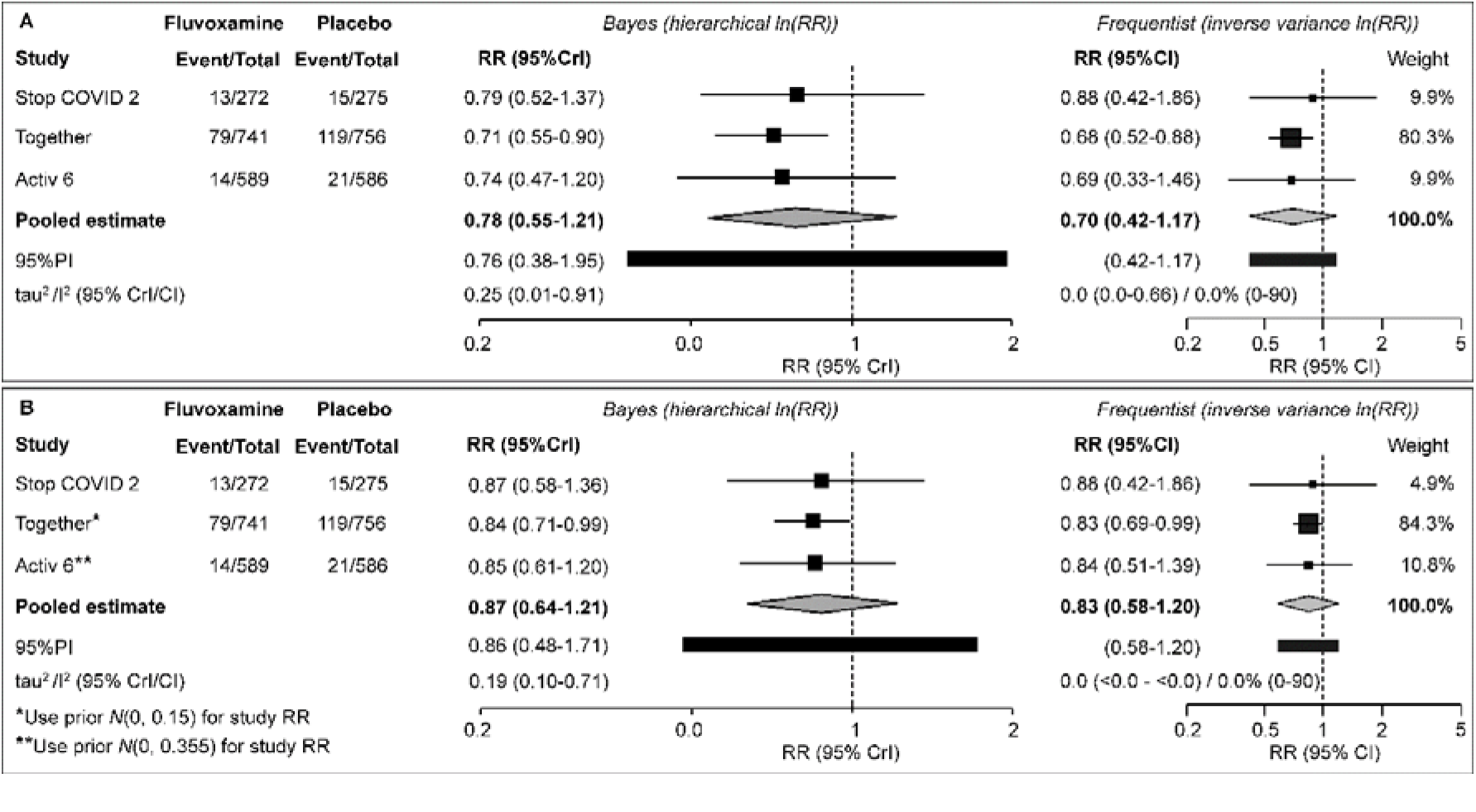
Random-effects meta-analysis of incidence of composite endpoints illustrating *disease deterioration* in three larger trials: Stop COVID 2 (3), Together (4) and Activ 6 (6). In the latter trial, this composite comprised unabigous components, while in the other two, elements susceptible to error and bias were incorporated (Table 2, Table 3). Meta-analyses were performed on reported (A), and on “corrected” (B) individual study effect estimates. Bayesian meta-analysis employed a moderately informed skeptical normal prior for the pooled estimate (*N*(0, 0.355) - fixed at 0 for ln(RR) with SD 0.355, assigns 95% probability to RRs 0.5-2.0, and weakly informative prior for τ^2^ (Cauchy (0, 0.5)). Study-level estimates are shrinkage estimates. Frequentist model was a generic inverse variance model with restricted maximum likelihood estimator with Hartung-Knapp-Siddik-Jonkman and ad-hoc variance correction. **A.** Meta-analysis of the reported individual study effect estimates. **B**. Meta-analysis of “corrected” individual study effect estimates: skeptical prior *N*(0, 0.15) (fixed at 0 for ln(RR) with standard deviation 0.15) instead of a flat one was used to cacluate relative risk in order to account for the interim analyses in the Together trial, and *N*(0, 0.355) (instead of flat) to correct RR in Activ 6, because originally this outcome was not intended for inferential purposes and no measures to control false-positive rate under multiple outcomes were implemented.

**Figure 4.**
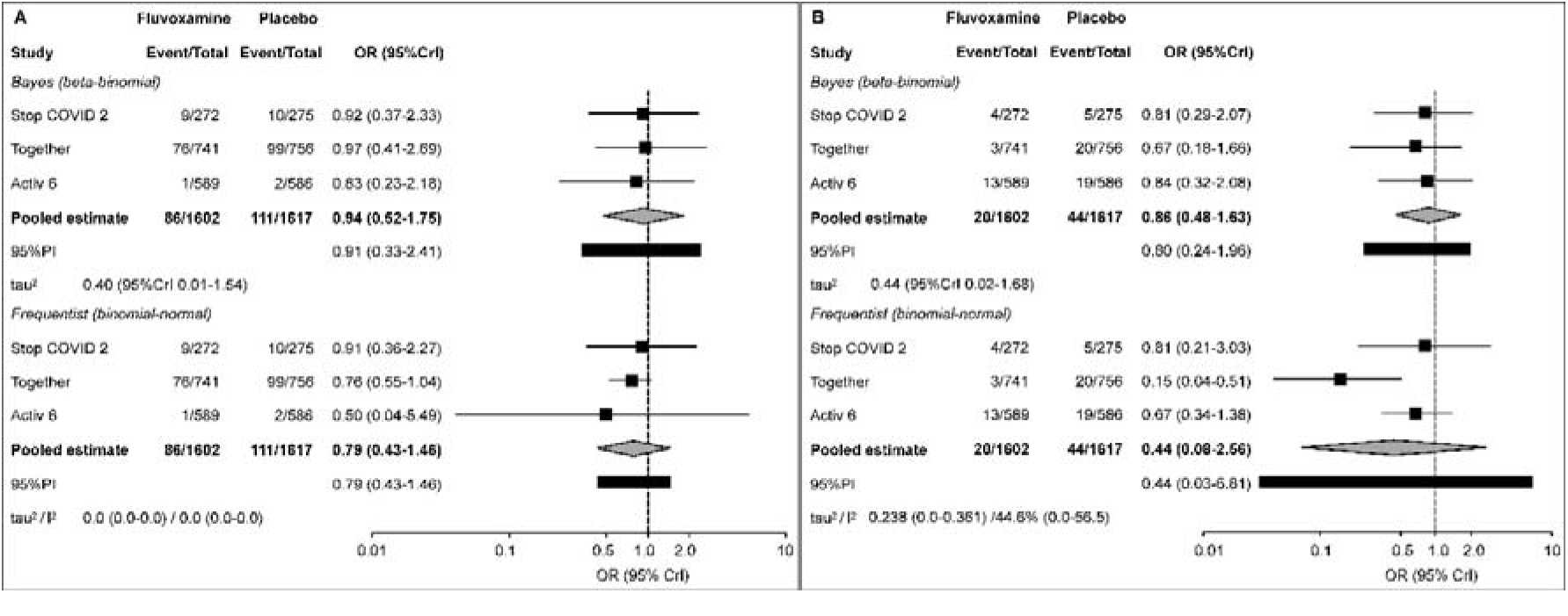
Random-effects meta-analysis of the reported raw incidence of hospitalization (**A**) and of other elements (cumulative “non-hospitalization” events) of the *deterioration* composites (**B**) in three larger trials: Stop COVID 2 (3), Together (4) and Activ 6 (6). Bayesian meta-analyses employed a moderately informed skeptical normal prior for the pooled estimate (*N*(0, 0.355) - fixed at 0 for ln(OR) with SD 0.355, assigns 95% probability to ORs 0.5-2.0, and weakly informative prior for τ^2^ (Cauchy (0, 0.5)). Study-level estimates are shrinkage estimates. Frequentist beta-binomial models did not converge, hence binomial-normal conditional model with approximate likelihood and t-distribution was fitted. Study-level estimates are model-generated estimates.

Even if one were to combine smaller and larger trials and assume no bias, based on the reported hospitalization frequencies, uncertainty regarding any benefit remains large (Figure 5): (i) Bayesian models (skeptical prior *N*(0,0.355) or uninformative prior *N*(0,1)) reveal substantial heterogeneity and wide prediction intervals, with pooled estimates (OR = 0.81, 95% CrI 0.48– 1.43, and OR = 0.63, 95% CrI 0.27–1.54, respectively) consistent with markedly lower or higher odds of hospitalization; (ii) frequentist models fail to estimate τ^2^ appropriately, resulting in too narrow confidence intervals, yet show no clear benefit (OR = 0.72, 95% CI 0.51–1.02).

**Figure 5.**
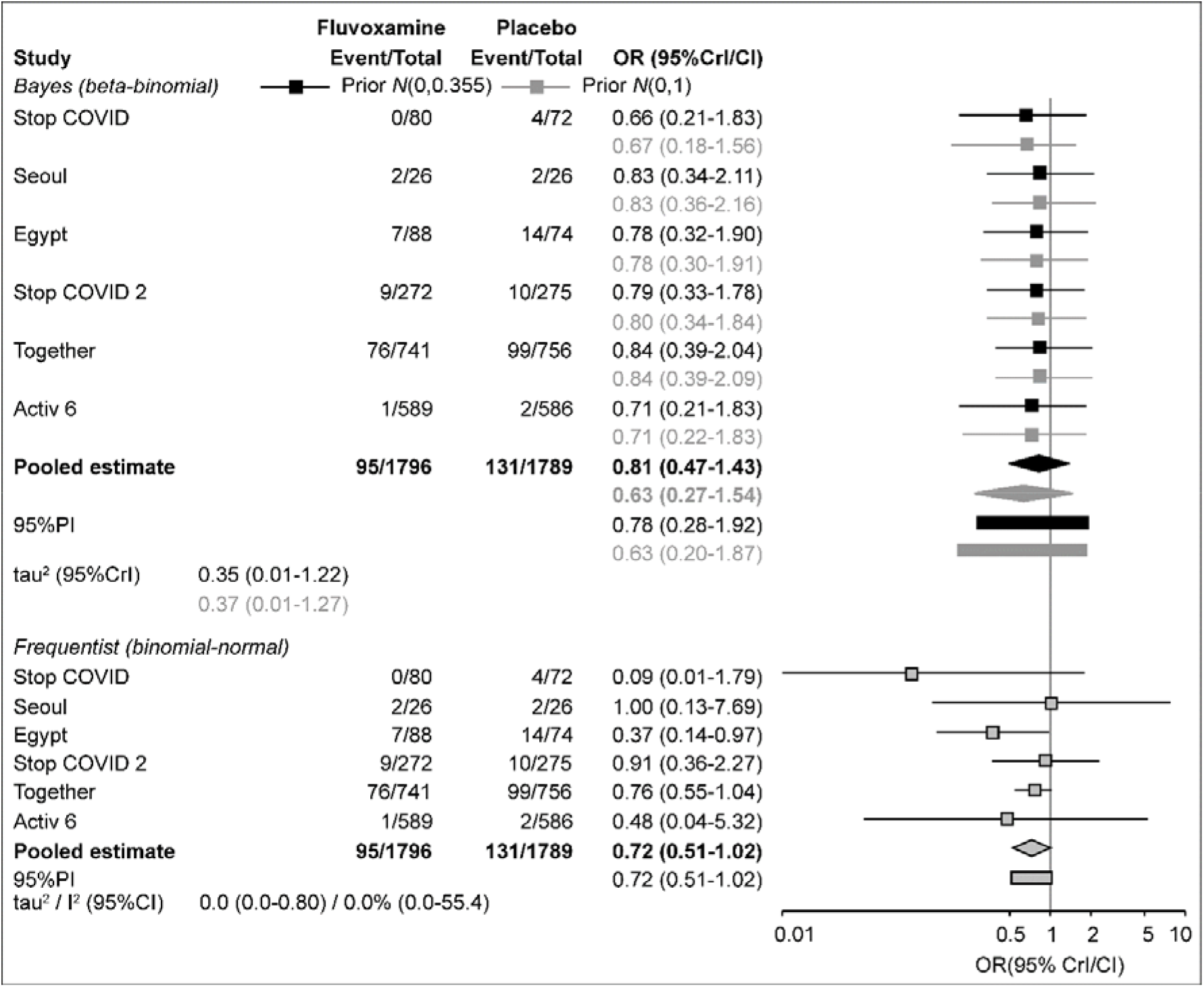
Random-effects meta-analysis of the reported raw incidence of hospitalization combining smaller and larger trials. Bayesian meta-analyses employed a moderately informed skeptical normal prior for the pooled estimate (*N*(0, 0.355) - fixed at 0 for ln(OR) with SD 0.355, assigns 95% probability to ORs 0.5-2.0, and an uninformative prior (*N*(0, 1) - fixed at 0 for ln(OR) with SD 1, assigns 95% probability to ORs 0.14-7.39. Prior for τ^2^ was weakly informative (Cauchy (0, 0.5)). Bayesian study-level estimates are shrinkage estimates. Frequentist model was a binomial-normal conditional model with approximate likelihood and t-distribution. Study-level estimates are model-generated estimates.

If only trials in unvaccinated patients are considered (Table 1)—Stop COVID, Stop COVID 2, Together, and the trial in Seoul—heterogeneity increases and widens uncertainty further: (i) there were 87/1119 events in fluvoxamine-treated patients and 115/1129 events in placebo patients; (ii) Bayesian analysis with a skeptical prior yields τ^2^ = 0.48 (95% CrI 0.01–1.80), pooled estimate OR = 0.87 (95% CrI 0.49–1.58), and 95% prediction OR = 0.82 (95% CrI 0.20– 2.24); (iii) analysis with an uninformative prior yields τ^2^ = 0.52 (95% CrI 0.02–1.98), pooled OR = 0.68 (95% CrI 0.23–1.97), and prediction OR = 0.68 (95% CrI 0.14–2.33); (iv) frequentist analysis yields τ^2^ = 0.0 (95% CI 0.0–5.58), I^2^ = 0.0% (95% CI 0.0–75.1), and pooled OR = 0.77 (95% CI 0.49–1.21).

Given six RCTs—still too few to explore heterogeneity thoroughly—and no obvious clinical or methodological marker (Tables 1 and 2) that consistently explains it, residual bias or chance is a more plausible explanation of the reported results and their heterogeneity than a true, context-dependent treatment effect.

## Discussion

This review was motivated by two recent meta-analyses of RCTs (or studies declared as such) claiming efficacy of “higher-dose” fluvoxamine in prevention of disease deterioration/hospitalization in adult outpatients with mild-to-moderate COVID-19 disease (8,9). Both were published after the FDA’s decision not to grant emergency use authorization for fluvoxamine in this indication (1), and no subsequent RCT has strengthened the “case for fluvoxamine.” The present review explicitly demonstrates why such claims are not substantiated by the primary studies on which they were based. From a practical standpoint, current claims about fluvoxamine—whether in support or opposition—are of limited clinical relevance. Nevertheless, unopposed assertions of efficacy could be misinterpreted as justified, potentially leading to inappropriate use. Beyond the specific case of fluvoxamine, this review emphasizes two broader points: (i) RCTs are complex models—all aspects of design, conduct, and analysis must be carefully considered when interpreting reported results; (ii) systematic reviews and meta-analyses are commonly undertaken superficially and are published indiscriminately (51). As Olkin notes (52, p. 299), *“A meta-analysis is not to be undertaken lightly; it is not a simple procedure, nor is it a cure-all. It is time-consuming, requires specialists knowledgeable about the treatment working with statisticians, and is subject to abuse; treat it with respect*.*”*

It could be objected that the present review was biased against fluvoxamine because Bayesian analyses used a moderately informed skeptical prior. This concern is mitigated by several observations: (i) frequentist estimates, generally appropriate for the data but limited regarding τ^2^ estimation, also failed to support efficacy claims; (ii) the skeptical prior used (N(0, 0.355)) for ln(OR) or ln(RR) is not overtly conservative—it assigns 95% probability to ORs/RRs between 0.5 and 2.0, i.e., it is “benevolent” toward marked benefits (e.g., 30–50% RRR); (iii) analysis using an uninformative prior yielded results closely similar to those obtained with the skeptical prior (Figure 5); (iv) in the context of Bayesian learning in medicine (53), a skeptical prior is consistent with pre-existing knowledge—although preclinical data suggested possible antiviral/anti-inflammatory effects (24), the grounds for an expected clinical benefit were weak. The prior therefore reflects reasonable starting assumptions while still permitting detection of substantial effects.

Under such conditions, the starting point is always the same: one wants to accurately quantify an effect, determine how far it is from “zero,” and assess how certain one can be about this distance. Proper quantification and communication of uncertainty about a treatment effect is crucial for its clinical relevance (54). In meta-analyses, uncertainty arises primarily from heterogeneity among individual study results (55). This review demonstrates substantial heterogeneity in the presumed effect of fluvoxamine on COVID-19 deterioration, which cannot be explained by patient characteristics, dosing, or treatment duration. Such heterogeneity is most likely attributable to biases in the design and conduct of the primary studies (56). Notably, the studies that suggested the strongest benefits—Stop COVID (2), the Egyptian trial (21), and the Together trial (4)—also exhibited the greatest susceptibility to bias or chance findings. These included issues in outcome definition, potential unblinding, statistical analysis, randomization, placebo adequacy, and sample size (Table 1, Table 2).

In conclusion, current randomized placebo-controlled trials do not support efficacy of any of the tested doses of fluvoxamine in prevention of disease deterioration in adult outpatients with mild-moderate COVID-19 disease. Claims to the contrary are not substantiated by the primary data, and the observed heterogeneity of the reported effects is more plausibly explained by bias and chance rather than true treatment benefit.

## Statements and declarations

### Data availabitliy

All data used in this work are available from the author on a reasonable request, however, all data are explicitly shown in tables and figures.

## References

1. Memorandum explaining basis for declining request for emergency use authorization of fluvoxamine maleate. US Food & Drug Administration website. Accessed February 20, 2026. Availabe at EUA 110 Fluvoxamine Decisional Memo_Redacted.pdf

2. Lenze EJ, Mattar C, Forumski CF, Stevens A et al. Fluvoxamine vs placebo and clinical deterioration in outpatients with symptomatic COVID-19. A randomized clinical trial. JAMA. 2020: 324(22):2292–2300. doi:10.1001/jama.2020.22760

3. Reiersen AM, Mattar C, Bender Ignacio RA et al. The Stop COVID 2 study: fluvoxamine vs placebo for outpatients with symptomatic COVID-19, a fully remote randomized controlled trial. Open Forum Infect Dis. 2023; 2023(10):ofad419. doi: 10.1093/ofid/ofad419

4. Reis G, dos Santos Moreira-Silva EA, Medeiros Silva DC et al. Effect of early treatment with fluvoxamine on risk of emergeny care and hospitalization among patients with COVID-19: the Together randomized, platform clinical trial. Lancet Glob Health. 2022; 10:e–42–51. 10.1016/S2214-109X(21)00448-4

5. Bramante CT, Juling JD, Tignanelli CJ et al. Randomized trial of metformin, ivermectin, and fluvoxamine for Covid-19. N Engl J Med. 2022; 387:599–619. doi: 0.1056/NEJMoa2201662

6. Stewart TG, Rebolledo PA, Mourad A et al. Higher-dose fluvoxamine and time to sustained recovery in outpatients with COVID-19. The ACTIV 6 randmized trial. JAMA. 2023; 330(24):2354–2363. doi:10.1001/jama.2023.23363

7. McCarthy MW, Naggie S, Boulware DR et al. Effect of fluvoxamine vs Placebo on time to sustained recovery in outpatients with mild to moderate COVID-19. A randomized clinical trial. JAMA. 2023; 329(4):296–305. doi:10.1001/jama.2022.24100

8. Zhou Q, Zhao G, Pan Y, Zhang Y, Ni Y. The efficacy and safety of fluvoxamine in patients with COVID-19: a systematic review and meta-analysis form randomized controlled trials. PloS ONE. 2024; 2024(19): e0300512. 10.1371/journal.pone.0300512

9. Prasanth MI, Leshan Wannigama, Riersen AM et al. A systematic review and meta-analysis investigating dose and time of fluvoxamine treatment efficacy for COVID-19 clinical deterioration, death and long-COVID complications. Sci Reports. 2024; 14:13462. 10.1038/s41598-024-64260-9

10. Crockar TF, Lam N, Jordao M et al. Risk-of-bias assessment using Cochrane’s revised tool for randomized trials (RoB 2) was useful but challenging and resource-intensive: observations from a systematic review. J Clin Epidemiol. 2023; 161: 39–45. doi:10.1016/j.jclinepi.2023.06.015

11. Kanukula R, McKenzie JE, Cashin AG et al. Variation observed in consensus judgments between pairs of reviewers when assessing the risk of bias due to missing evidence in a sample of published meta-analyses in nutrition research. J Clin Epidemiol. 2024; 166: 111244 10.1016/j.jclinepi.2023.111244

12. Mathur MB, VanderWeele TJ. Methods to address confounding and other biases in meta-analysis: review and recommendations. Annu Rev Public Health. 2022; 43:19–35. doi: 10.1146/annurev-publhealth-051920-114020

13. Brooks ME, Kristensen K, van Benthem KJ et al. GlmmTMB balances speed and flexibility among packages for zero-inflated generalized linear mixed modeling.The R Journal. 2017; 9(2): 378–400. doi:10.32614/RJ-2017-066

14. Viechtbauer W. Conducting meta-analyses in R with the matagor package. J Stat Software. 2010; 36(3): 1–48. 10.18637/jss.v036.i03

15. Balduzzi S, Rucker G, Schwarzer G. How to perofrm a meta-analysis with R: a practical tutorial. Evid Based Ment Health. 2019; 22(4):153–160. doi: 10.1136/ebmental-2019-300117

16. Burkner PC. Brms: an R package for Bayesian multilevel models using Stan. J Stat Software 2017; 80(1). 10.18637/jss.v080.i01

17. Siripongboonsitti T, Ungtrakul T, Tawinprai K et al. Efficacy of combination therapy of fluvoxamine and favipiravir vs favipiravir monotherapy to prevent severe COVID-19 among mild to moderate COVID-19 patients: open-label randomized controlled trial (EFFaCo study). Int J Infect Dis. 2023; 134:211–219. doi: 10.1016/j.ijid.2023.06.018

18. Reis G, dos Santos Moreira Silva, Medeiros Silva DC et al. Oral fluvoxamine with inhaled budesonide for treatment of early-onset COVID-19. Ann Intern Med. 2023; 176(5):667–675. doi:10.7326/M22-3305

19. Reis G, dos Santos Moreira Silva EA, Medeiros Silva DC et al A multi-center, adaptive, randomized, platform trial to evaluate the effect of repurposed medicines in outpatients with early coronavirus disease 2019 (COVID-19) and high-risk for complications: the TOGETHER master trial protocol. Gates Open Res. 2021; 5:117 10.12688/gatesopenres.13304.2

20. Seo H, Kim H, Bae S et al. Fluvoxamine treatment of patients with symptomatic COVID-19 in a community treatment center: a preliminary result of randomized controlled trial. Infect Chemother. 2022; 54(2):102–113. doi: 10.3947/ic.2021.0142

21. Ibrahim MA, Shehta M. Treatment with fluvoxamine in nonhispitalized coronavirus disease 2019 patients. Egypt J Chest Dis Tuberculosis. 2023; 72:40–45. doi: 10.4103/ecdt.ecdt_38_22

22. Wannigama DL, Hurst C, Phattharapornjaroen P et al. Early treatment with fluvoxamine, bromhexine, cyproheptadine, and niclosamide to prevent clinical deterioration in patients with symptomatic COVID-19: a randomized clinical trial. EClinicalMedicine. 2024; 70:102517. 10.1016/j.eclinm.2024.102517

23. Dodds MG, Doyle EB, Reiersen AM, Brown F, Ranyer CR. Fluvoxamine for the treatment of COVID-19. Lancet Glob Health. 2022; 2022(10):e332. 10.1016/S2214-109X(22)00006-7

24. Sukhatme VP, Reiersen AM, Vayttaden SJ, Sukhatme VV. Fluvoxamine: a review of its mechanisms of action and its role in COVID-19. Front Pharmacol. 2021; 12:652688 10.3389/fphar.2021.652688

25. Pearson T, Caporaso JG, Yellowhair M et al. Effects of ursodeoxycholic acid on the gut microbiome and colorectal adenoma development. Cancer Med. 2019; 8(2):617–628. doi: 10.1002/cam4.1965

26. FDA warns about limitations and accuracy of pulse oximeters. US Food & Drug Administration website. Accessed February 20, 2026. Available at: FDA warns about limitations and accuracy of pulse oximeters

27. Ryan EG, Brok K, Gates S, Slade D. Do we need to adjust for interim analyses in a Bayesian adaptive trial design? BMC Res Methods. 2020; 20:150. 10.1186/s12874-020-01042-7

28. Senn S. Seven myths of randomization in clinical trials. Stat Med. 2013; 32(9):1439–1450. doi: 10.1002/sim.5713

29. Kernan WN, Viscoli CM, Makuch RW, Brass LM, Horwitz RI. Stratified randomization for clinical trials. J Clin Epidemiol. 1999; 52(1):19–26. doi: 10.1016/s0895-4356(98)00138-3

30. Guideline on adjustment for baseline covariates in clinical trials. EMA/CHMP/295050/2013. Accessed February 20, 2026. Available at: https://www.ema.europa.eu/en/documents/scientific-guideline/guideline-adjustment-baseline-covariates-clinical-trials_en.pdf

31. Hammond J, Leister-Tebbe H, Gardner A et al. Oral nirmatrelvis for high-risk, nonhospitalized adults with Covid-19. N Engl J Med. 2022; 386(15): 1397–1408. doi: 10.1056/NEJMoa2118542

32. Tierney JF, Burdett S, Fisher DJ. Practical methods for incorporating summary time-to-event data into meta-analysis: updated guidance. Syst Rev. 2025; 13:84 10.1186/s13643-025-02752-z

33. Schneeweiss S. Sensitivity analysis and external adjustment for unmeasured confounders in epidemiologic database studies. Pharmacoepidemiol Drug Saf. 2006; 15(5):291–303. doi: 10.1002/pds.1200

34. Hamid S, Derado G, Pham H et al. Chronic conditions as risk factors for COVID-19-associated hospitalizations among adults 2020-2023. Am J Prevent Med. 2026; 2026(70):108227 10.1016/j.amepre.2025.108227

35. Inada-Kim M, Chmiel FP, Boniface M et al. Validation of oxygen saturations measured in the community by emergency medical services as a marker of clinical deterioration in patients with confirmed COVID-19: a retrospective cohort study. BMJ Open. 2024; 14: e067378. doi:10.1136/bmjopen-2022-067378

36. Boutron I, Page MJ, Higgins JPT et al. Chapter 7. Considering bias and conflicts of interest among the included studies. In: Higgins JP, Thomas J, Chandler J et al., eds. Cochrane handbook for systematic reviews of interventions version 6.5. Cochrane; 2024. Accessed February 15, 2026. Available at Chapter 7: Considering bias and conflicts of interest among the included studies | Cochrane

37. Savović J, Turner RM, Dawdsley D et al. Association between risk-of-bias assessment and results of randomized trials in Cochrane reviews: the ROBES meta-epidemologic study. Am J Epidemiol 2018; 187(5): 1113–1122. doi: 10.1093/aje/kwx344

38. Aung NM, Jurak I, Mehmood S, Axon E. Sensitivity analysis in meta-analysis: a tutorial. Cochrane Evid Synth Methods. 2026; 2026(4):e70067 10.1002/cesm.70067

39. Langan D, Higgins JPT, Jackson D et al. A comparison of heterogeneity variance estimators in simulated random-effects meta-analyses. Res Synth Methods. 2018; 10(1): 83–98. doi: 10.1002/jrsm.1316

40. Pateras K, Nikolakopoulos S, Mavridis D, Roes KCB. Interval estimation of the overall treatment effect in a meta-analysis of a few small studies with zero events. Contemp Clin Trials Commun. 2018; 9:98–107. doi: 10.1016/j.conctc.2017.11.012

41. Yao M, Deng K, Wang Y, Mei F, Zou K, Li L et al. Random-effects meta-analysis models for pooling rare events data: a comparison between frequentis and Bayesian methods. BMC Res Method. 2025; 25:228 10.1186/s12874-025-02664-5

42. Kuss O. Statistical methods for meta-analyses including information from studies without any events – add nothing to nothing and succeed nevertheless. Stat Med. 2015; 34(7):1097–1116. doi: 10.1002/sim.6383

43. Stijnen T, Hamza TH, Ozdemir P. Random effects meta-analysis of event outcome in the framework of the generalized linear mixed model with applications in sparse data. Stat Med. 2010; 29(29):3046–3067. doi: 10.1002/sim.4040

44. Friede T, Rover C, Wander S, Neuenschwnder B. Meta-analysis of fes small studies in orphan disease. Res Synth Method. 2017; 8(1):79–91. doi: 10.1002/jrsm.1217

45. Sterne JA, Gavaghan D, Egger M. Publication and related bias in meta-analysis: power of statistical test and prevalence in the literature. J Clin Epidemiol. 2000; 53(11):1119–1129. doi: 10.1016/s0895-4356(00)00242-0

46. Deschartres A, Trinquart L, Boutron I, Ravaud P. Influence of trial sample size on treatment effect estimates: meta-epidemiological study. BMJ. 2013; 346: f2304 doi: 10.1136/bmj.f2304

47. IntHout J, Iaonnidis JPA, Borm GF, Geoman JJ. Smalls studies are more heterogeneous than large ones: a meta-meta-analysis. J Clin Epidemiol. 2015; 68(8):860–869. doi: 10.1016/j.jclinepi.2015.03.017

48. Higgins JP, Li T, Deeks JJ (editors). Chapter 6. Choosing effect measures and computing estimates of effect. In: Higgins JP, Thomas J, Chandler J et al., eds. Cochrane handbook for systematic reviews of interventions version 6.5. Cochrane; 2024. Accessed February 15, 2026. Available at Chapter 6: Choosing effect measures and computing estimates of effect | Cochrane

49. VanderWeele TJ. Optimal approximate conversios of odds ratios and hazard ratios to risk ratios. Biometrics. 2020; 76(3):746–752. doi: 10.1111/biom.13197

50. Morea MS, Nestoriuc Y, Rief W. Lessons learned from placebo groups in antidepressant trials. Philos Trans R Soc Long B Biol Sci. 2011; 366(1572):1879–1888. doi: 10.1098/rstb.2010.0394

51. Ioannidis JPA. The mass production of redundant, misleading, and conflicted systematic reviews and meta-analyses. Milbank Q. 2016; 94(3):485–514. doi: 10.1111/1468-0009.12210

52. Olkin I. Invited commentary: Re:”A criticallook at some popular meta-analytic methods”. Am J Epidemiol. 1994; 140(3):297–299. doi: 10.1093/oxfordjournals.aje.a117249

53. Gill JC, Sabin L, Schmid CH. Why clinicians are natural Bayesians. BMJ. 2005; 330(7499):1080–1083. doi: 10.1136/bmj.330.7499.1080

54. Guyatt GH, Oxman AD, Vist GE, Kunz R, Flack-Ytter Y, Schunemann HJ. GRADE: what is “quality of evidence” and why is it important to clinicians. BMJ. 336(7651):995–998. doi: 10.1136/bmj.39490.551019.BE.

55. Guyatt GH, Oxman AD, Kunz R et al. GRADE guidelines: 7. Rating the quality of evidence – inconsistency. J Clin Epidemiol. 2011; 64(12):1294–1302. doi: 10.1016/j.jclinepi.2011.03.017

56. Rhodes KM, Turner RM, Jones HE, Mawdsley D, Higgins JPT. Between-trial heterogeneity in meta-analyses may be partly explained by reported design characteristics. J Clin Epidemiol. 2018; 95:45–54. doi: 10.1016/j.jclinepi.2017.11.025

